# Underdiagnosis Bias of Chest Radiograph Diagnostic AI can be Decomposed and Mitigated via Dataset Bias Attributions

**DOI:** 10.1101/2024.12.16.24319063

**Authors:** Yuta Kobayashi, Haoran Zhang, Harvineet Singh, Judy Wawira Gichoya, Shalmali Joshi

## Abstract

Inequitable diagnostic accuracy is a broad concern in AI-based models. However, current characterizations of bias are narrow, and fail to account for systematic bias in upstream data-collection, thereby conflating observed inequities in AI performance with biases due to distributional differences in the dataset itself. This gap has broad implications, resulting in ineffective bias-mitigation strategies. We introduce a novel retrospective model evaluation procedure that identifies and characterizes the contribution of distributional differences across protected groups that explain population-level diagnostic disparities. Across three large-scale chest radiography datasets, we consistently find that distributional differences in age and confounding image attributes (such as pathology type and size) contribute to poorer model performance across racial subgroups. By systematically attributing observed underdiagnosis bias to distributional differences due to biases in the data-acquisition process, or dataset biases, we present a general approach to disentangling how different types of dataset biases interact and compound to create observable AI performance disparities. Our method is actionable to aid the design of targeted interventions that recalibrate foundation models to specific subpopulations, as opposed to methods that ignore systematic contributions of upstream data biases on inequitable AI performance.

## INTRODUCTION

There is growing concern that AI algorithms deployed in sensitive domains may replicate or exacerbate inequities in diagnosis or patient outcomes^1^. Current characterizations of bias of AI performance conflates observed retrospective disparities with algorithmic inequity without accounting for or characterizing the impact of upstream biases contributing to biased datasets. Broadly, this has resulted in fairness interventions that are not generalizable, requiring unjustified trade-offs, further disconnecting the impact of broader systemic disparities and the resulting biases in observational training data with that of inequities in AI performance^2,3^. Addressing algorithmic inequities requires a shift from the narrow focus on retrospective model performance to characterizing the impact of systematic upstream biases in the training data-generating processes. To that end, we propose to characterize the contribution of various granular distributional differences of patient characteristics across subpopulations within a dataset, to disentangle the causes of AI performance differences, arising from (a combination of) differences in data acquisition procedures or clinically relevant disease-related differences among subgroups. While our approach is general, we operationalize this auditing pipeline on evaluating biased performance of foundation models of disease diagnosis from chest radiology images^4-7^.

While chest radiography-based AI diagnostic algorithms may match specialist performance^8^, with clear potential for real-world deployment^9^, there is consistent evidence of population-level performance disparity, particularly underdiagnosis among underrepresented groups in public datasets^5^. Underdiagnosis bias may have significant ramifications such as causing delayed access to care for historically under-served minority populations^1,10,11^. In X-ray disease detection, the AI algorithm is often trained and evaluated agnostic to important knowledge of the population demographics, patient characteristics (such as disease prevalence differences), clinical workflow, and operational factors constituting the upstream data-acquisition process. These upstream mechanisms may influence the patterns learned by the AI model, even if an AI model does not explicitly rely on these variables during training. For example, differences in healthcare utilization patterns may lead to differences in the image characteristics and manifestation of pleural effusion across demographic subpopulations, leading to model performance degradation for underrepresented patients.

We perform a systematic study of underdiagnosis rates across racial subgroups in AI-based deep learning and foundation models on three chest X-ray datasets (MIMIC-CXR, CheXpert, Emory). To do so, we use a novel decomposition of the observed performance disparity to separate components representing different types of dataset biases^12,13,14^. We apply the method by Zhang et. al.^15^ to assess the relative contribution of error disparity from different sources such as shifts in population demographics, clinical workflow (such as views collected *conditioned on initial impression*), disease prevalence, or variability in the X-ray (*conditioned on demographics, prevalence, and workflow variability*) a model is sensitive to. Our method is more reliable and actionable since it accounts for the complexity of the data-generating process as opposed to canonical approaches to interpreting and explaining AI behavior^16-18^. Our method is able to surface consistent and multifactorial causes of AI diagnostic disparities across all datasets. We further show how our attributions can be used to inform targeted model adaptations. Our analysis highlights that extraction and rigorous characterization of the impact of upstream distributional differences on model performance, beyond knowledge of patient demographics, is imperative for advancing the understanding of algorithmic bias and the development of reliable and generalizable bias-mitigation strategies.

## RESULTS

### Disease Diagnosis Tasks and Datasets

We focus on two performance metrics based on prior work^5^ that assess the performance of two distinct tasks; one task aims to rule out the presence of any disease, and a second task aims to detect a specific pathology (pleural effusion). Pleural effusion was chosen as a pathology of interest since previous studies have reported disparate performance^4^. For the first task, the performance metric of interest is the underdiagnosis rate, defined as the False Positive Rates (FPR) for the binarized model prediction for the ‘No Finding’ label^5^, indicating that no disease is diagnosed from the patient chest X-ray image. For the second task, we define the pathology detection rate as the True Positive Rate (TPR) for the ‘Pleural Effusion’ label. We choose to study the diagnostic disparities between White and Black patients, given that Black patients are sufficiently represented across the three datasets and there are consistent observable performance disparities across tasks. Our analysis pipeline is summarized in Fig. 1.

**Figure 1:**
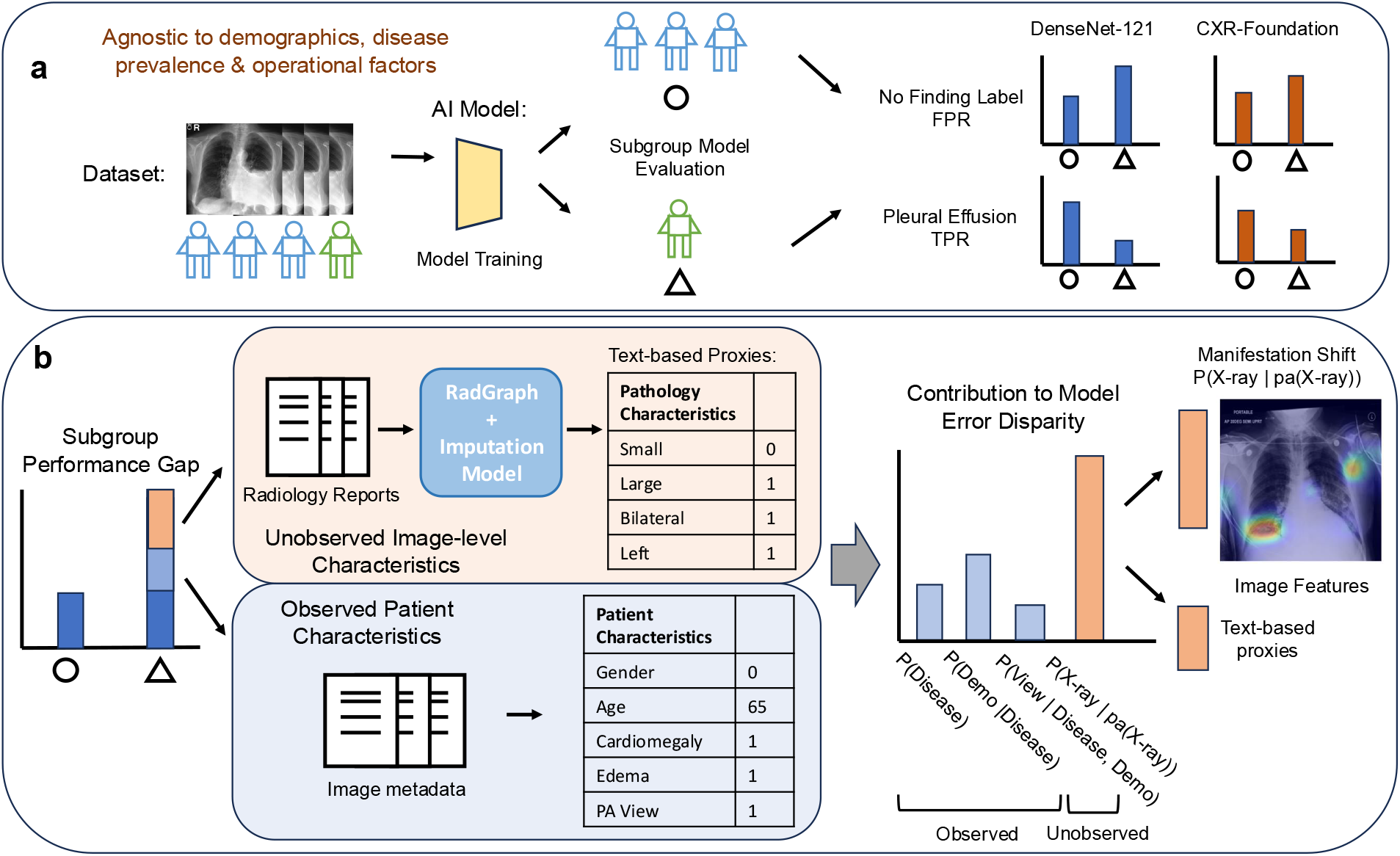
Overview of our analysis pipeline. **a.** We train distinct multi-label DenseNet-121 models for each dataset. We compute the subgroup-level model performance disparity using the underdiagnosis rate (FPR of the ‘No Finding’ label) and the pleural effusion detection rate (TPR of ‘Pleural Effusion’ label) for DenseNet-121 as well as the CXR-Foundation model. **b**. We decompose the total performance disparity to contributions of different confounding factors using our attribution method, both observed (differences in pathology types, demographics, image acquisition parameters from meta-data) and unobserved (differences in pathology manifestation and image features).

We perform our analysis using three datasets, MIMIC-CXR^19^, CheXpert^20^, and Emory-CXR^21^. Pathology labels were generated using CheXpert labeler^20^ across all datasets. We train a DenseNet-121 neural network to classify 14 different conditions using multi-task learning, using only the chest X-rays as input (Fig 1a.,1b.). Each dataset was randomly split using a ratio of 60-10-30 for train-validation-test, and a separate model was trained on each dataset. We follow the same architecture and training procedure as previous work^5^, detailed in the Methods. We also analyze a state-of-the-art foundation model from Google^22^ pre-trained on a completely distinct set of weakly labeled chest X-rays from USA and India, called CXR-foundation. Note that we cannot analyze CXR-foundation on Emory data due to privacy concerns. We report the final demographic distribution of the evaluation cohort used in the diagnostic disparity analysis for each dataset in Table 1.

**Table 1.**
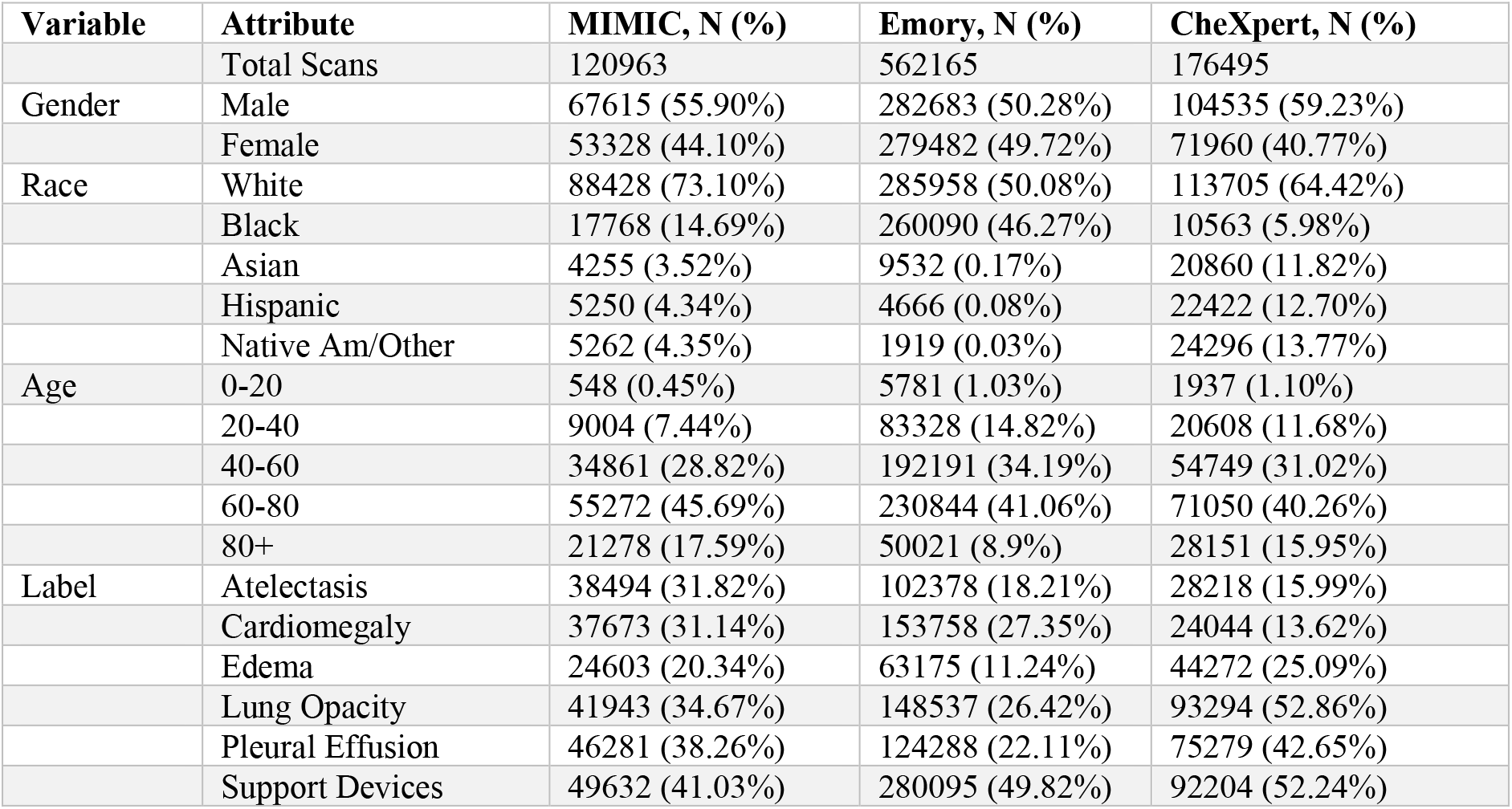
Summary statistics for the evaluation cohort used for the disparity analysis in each dataset. The cohort only includes patients with findings in the X-ray, and patients with missing demographics (gender, age, race) are dropped.

### Underdiagnosis Rates across Datasets and Models

We plot the mean and standard deviation for the model performance metrics of interest (“No Finding” FPR, “Pleural Effusion” TPR) in the evaluation cohort across datasets and racial subgroups for the DenseNet-121 and CXR-Foundation models (Fig. 2). The standard deviation for the model performance is obtained by bootstrapping the training dataset and retraining the final classification layer, while keeping the test dataset fixed. We find consistent patterns across datasets, where underrepresented subgroups are underdiagnosed at a higher rate (Fig. 2a). Additionally, there is a consistent pattern across datasets where Black patients have lower detection rates for pleural effusion (Fig. 2b).

**Figure 2:**
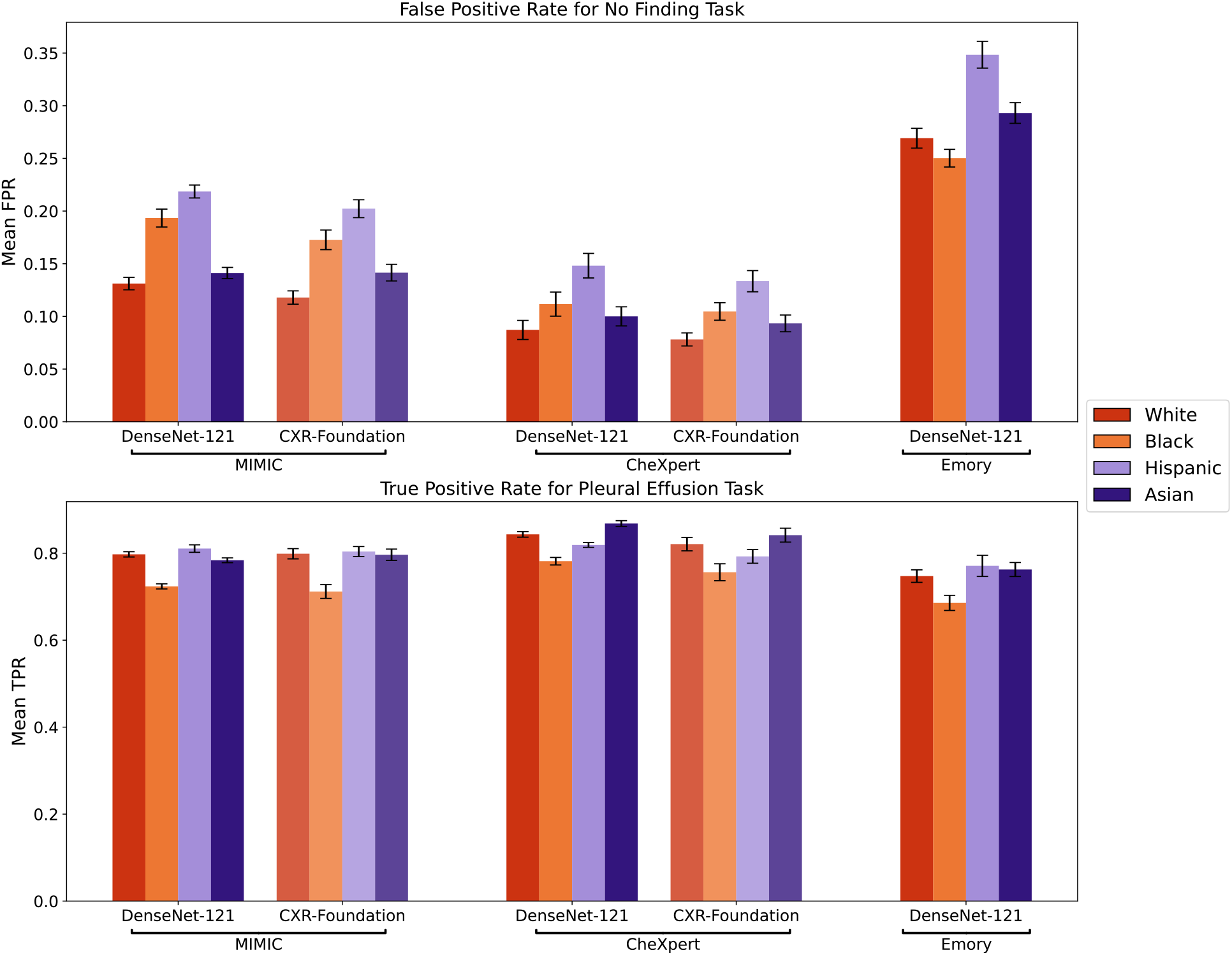
Racial performance disparities for “No Finding” and “Pleural Effusion” tasks across datasets for DenseNet-121 and CXR-Foundation Models, with error bars representing standard deviation across 20 bootstrap iterations. **a.** For MIMIC-CXR and CheXpert, the subgroup with the best performance is White patients. Underdiagnosis is most observed for Black and Hispanic patients. For Emory, the subgroup with the best performance is Black patients. Underdiagnosis occurs for Hispanic, White, and Asian patients. CXR-Foundation is better able to disambiguate healthy and diseased X-rays, as evident by consistently lower FPR rates. **b**. Model performance is consistently worse for Black patients for the task of detecting pleural effusion. The pattern of underdiagnosis is also consistent across models for MIMIC-CXR and CheXpert. A table with numerical values and statistical comparisons is included in the Supplement.

**Figure 3:**
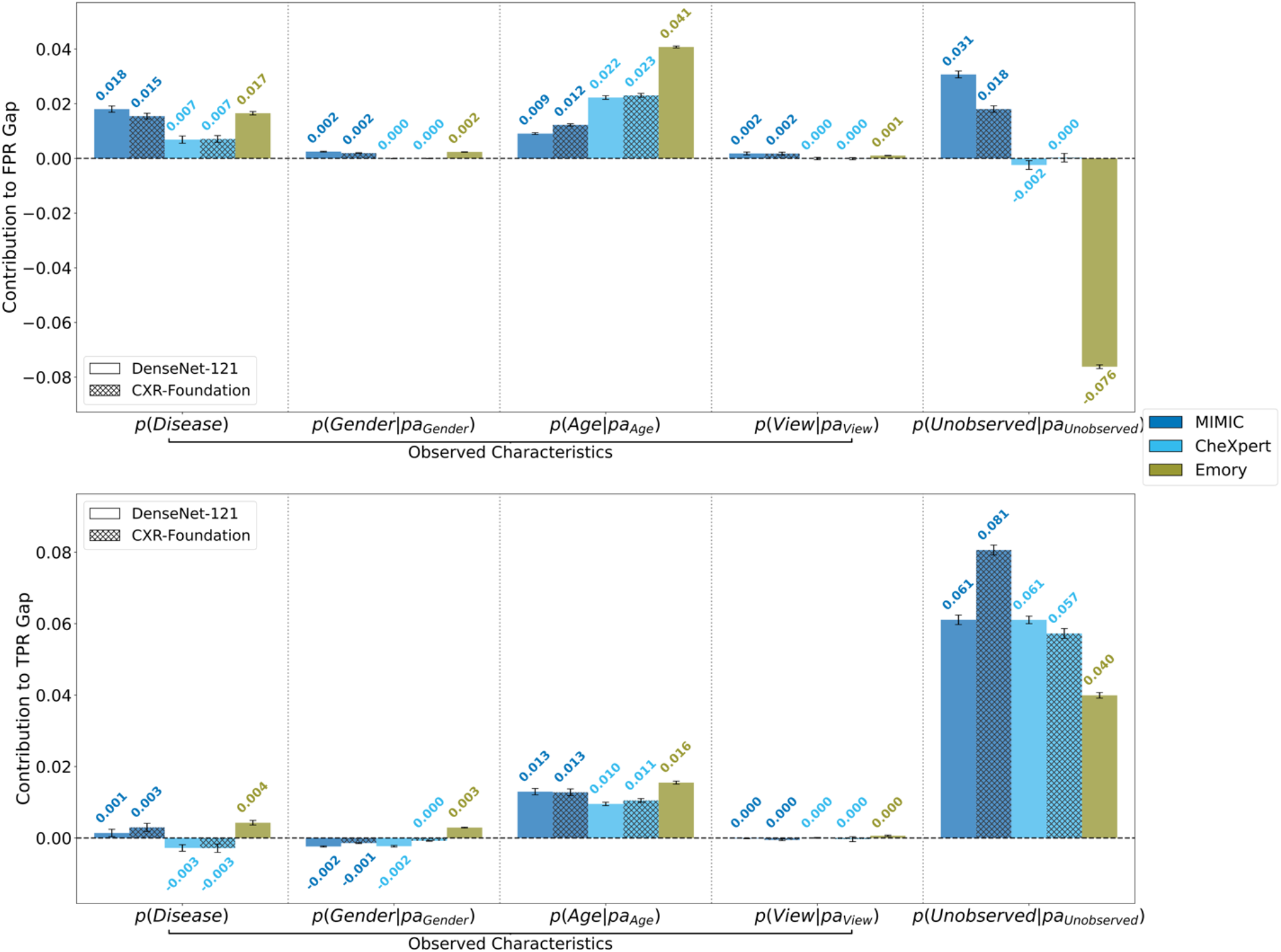
Contribution of each distribution shift mechanism to performance disparity across Black and White patients, error bars represent standard deviation across 100 bootstrap iterations. **a.** A larger positive attribution value (y-axis) indicates a larger contribution to the overall false positive rate (FPR) disparity for the ‘No Finding’ label. The x-axis shows the five different mechanisms that contribute to overall performance disparity. For example, a shift in *p*(*Age* |*pa*_*Age*_) indicates a shift in the age distribution given parent variables, which in this case are pathology and gender. Marginal disease prevalence shifts, and conditional age shifts are consistent contributors to greater underdiagnosis rates for Black patients. The difference in X-ray attribution across datasets can arise for various reasons, such as sample classification difficulty or label error. **b**. A larger positive attribution value (y-axis) indicates a larger contribution to the total true positive rate (TPR) disparity for the ‘Pleural Effusion’ label. Distributional differences of age result in lower detection rate among Black patients. However, observed patient characteristics do not fully explain the TPR disparity, indicating the presence of other unobserved or spurious features in the X-ray affecting model performance.

We note that CXR-Foundation performs uniformly better (Table S1) in the task of differentiating no-finding X-rays across all racial groups compared to the dataset-specific DenseNet-121 models. The performance disparities across racial groups are also smaller, highlighting the benefits of pretraining larger capacity models on large, diverse datasets. However, the patterns of underdiagnosis bias across racial groups persist, indicating that model capacity does not fully explain observable performance disparities. Similarly, lower detection rates of pleural effusion in Black patients can also be observed with CXR-Foundation, but patterns of diagnosis disparity persist across models and sites (Table S2).

### Decomposing Underdiagnosis Bias Across Patient Subgroups

We hypothesize that distributional differences in patient demographics and clinical characteristics obtained from meta-data, i.e., differences not observed by the model during training, partially explain observable error rate disparities across racial subgroups and enable reliable attribution of disparities in terms of these differences. Importantly, this is the case *even if our candidate models do not explicitly use these variables as input*.

For the observed patient confounding attributes, we consider the binary pathology labels *Z*, demographics *D*, and the view position of the X-ray *C*. These variables were selected as they are extracted consistently across all datasets and may causally influence the distribution of image features^23,24^. The demographics variable includes gender and age group all encoded as binary variables. The view position is represented using a binary indicator variable *C* ={*C*_*PA*_, *C*_*AP*_, *C*_*LA*_, . . . } for each view type (AP: anteroposterior, LA: lateral, etc.). The acquisition of various views is based on clinical guidelines, depending on the suspected pathology as some view positions allow for better detection or localization of pathologies like cardiomegaly and pneumothorax volume^25^. Demographic attributes may also influence the ability for patients to obtain specific X-ray views, due to being in a severe condition or being physically unable to position themselves for a PA view^26^.

We consider a source and target distribution corresponding to two patient subgroups, denoted by *P* and *Q* respectively. Our goal is to decompose the error rate disparity defined by the total difference in model performance metric *L* (such as FPR/TPR) into further granular and informative components.

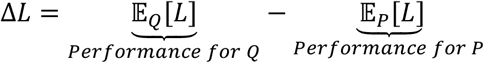

We attribute the performance gap to distributional differences in each aggregate factor (pathologies *Z*, demographics *D*, view-type *C*, and X-ray features *X*). We choose the following factorization of the joint probability distribution *p*(*Z*, *D*, *C*, *X*) based on knowledge of the data-generating process, as shown below.

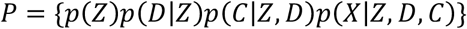

Each factor represents a probabilistic mechanism or dependence structure between variables that can shift across racial subgroups. In other words, the distribution shift in *p*(*Z*, *D*, *C*, *X*) from *P* to *Q* can be decomposed into four shifts; A shift in *p*(*Z*) represents a prevalence shift of pathologies across race (in *Q*(*Z*)). A shift in *p*(*D*|*Z*) indicates that for a certain disease, the age or gender representation may differ across race. A shift in *p*(*C*|*Z*, *D*) may arise as a shift in a clinician’s acquisition workflow for certain diseases or demographic characteristics, for instance if lateral views are obtained more often for one subgroup of patients. Finally, a shift in *p*(*X*|*Z*, *D*, *C*) may be indicative of manifestation shift given a certain age, gender, and pathology. Given that distribution shifts in image-level features are difficult to estimate and interpret, we aim to decompose this error contribution using text-based proxies as well as visualizing distribution shifts in the model’s feature representation space using saliency maps (Fig. 1b).

Note that each factor is a distribution conditioned on or adjusting for variables that influence the confounding variable according to the data-generating process. For example, we consider the conditional distribution *p*(*C*|*Z*, *D*) as the view position may be influenced by patient demographics and pathologies. This enables our attribution to reliably decompose error contributions across multiple confounding factors. A formalism of our approach based on causal models is provided in the Supplementary materials.

### Prevalence and Age Shifts Contribute to Diagnostic Disparity

We plot the results of our attribution method, where the x-axis indicates the distribution of each aggregate confounding factor. For each of these distributional factors, we plot the Shapley value on the y-axis. Each Shapley value represents the additive contribution of the factor to the increased FPR of the “No Finding” label in Black patients. For example, an attribution of 0.022 for ‘age’ indicates that the shift in the conditional distribution of age given gender and disease from White patients to Black patients lead to an increase in FPR of 2.2% for Black patients. Note the relative contribution can be negative, indicating that the distribution shift reduced the FPR gap.

We find that disease prevalence shifts consistently leads to underdiagnosis and unfavorable model performance for Black patients. Across all three datasets, White patients have a higher prevalence of pleural effusion, and Black patients have a higher prevalence for cardiomegaly (Extended Data Fig. 1-3). One plausible explanation is that for each disease, the relative task difficulty may differ inherently, leading to lower underdiagnosis rates in Whites.

Furthermore, distributional differences in age lead to underdiagnosis and an increase in FPR for Black patients, even after adjusting for disease prevalence. Interestingly, Black patients are consistently younger compared to White patients across all datasets (Extended Data Fig. 4). The model is systematically performing worse on younger patients, potentially due to differences in how the pathology manifests between younger and older patients. For example, the same pathology may be smaller in size and harder to detect in younger patients, if the condition is less severe compared to older patients. We note that the magnitude of disease prevalence and age attributions is similar for CXR-Foundation, indicating that the mechanisms for causing systematic differences in performances are agnostic to model capacity.

Lastly, while the X-ray attribution may be due to a combination of many factors such as insufficient model capacity, we hypothesize that one contributing factor may be due to different prevalences of label noise^27,28^ across racial subgroups. Given that the labeler is trained on radiology reports in the CheXpert dataset, it is possible that a higher frequency of ambiguous cases is being incorrectly labeled as “No Finding” (a false positive label) in MIMIC-CXR and Emory, leading to larger disparity attribution to unobserved factors. Conversely, low label noise in the CheXpert dataset may explain why there is no attribution to the unobserved factors. The distributions of predicted probabilities (Fig. S3) also suggest that MIMIC-CXR and Emory contain more false positive samples that are likely to be due to labeler error.

### Image-level Feature Shifts Influence Pathology Detection Rates

We plot the attributions for the detection rate disparity in pleural effusion, using the same method and subgroup configurations as the previous section. We find that age differences across race consistently lead to worse model performance among Black patients, indicating that both the DenseNet-121 and CXR-Foundation models are systematically failing to detect effusion in younger patients (Fig. S4). However, across all datasets the performance disparity is still largely unexplained after adjusting for differences in demographic and pathology composition across race, indicating the presence of additional manifestation shifts in the image-level features. For example, pathology manifestation differences including the location, size, and ambiguity of the pleural effusion in the X-ray may influence the ability for the model to detect the disease, but such disease-related characteristics are “unobserved” as they are not recorded as meta-data in the dataset (Fig. 1b).

To this end, we perform a sensitivity analysis using text-based proxies of hypothesized unobserved confounders extracted from radiology reports. Our results (Fig. S1) shows that this age attribution can be partially explained away by effusion characteristics such as size, location, and radiologist uncertainty. This indicates that the model may be influenced by a combination of purely spurious age-related features, as well as “mixed” features that mix the label signal (pleural effusion) and age, such as effusion size and severity. We include a discussion of shortcut learning^29,30,31^ and spurious features in the Supplement.

However, the performance disparity is still largely unexplained after correcting for distributional differences in our extracted proxy variables. We hypothesize that there may be types of effusion indications or image features that are underrepresented in the general population but encountered more often in the Black population. To perform attributions with respect to image features, we combine our framework with commonly used saliency methods^32,33^ to identify image-based features that explain the component of error disparity arising from manifestation shift. We observe that the predicted probability of effusion is relying on holistic information in the lung regions (Fig. 4). Furthermore, we observe some consistent patterns where the features that are most predictive of disparity are the lower left regions of the lung, potentially indicative of the absence of some indication of pleural effusion. However, spurious structures outside of the lungs such as bone density and structures in the central region may also be informative for the error disparity. Based on the saliency maps, an evaluation team could potentially discern why the model diagnosis rates may differ across these X-rays.

**Figure 4:**
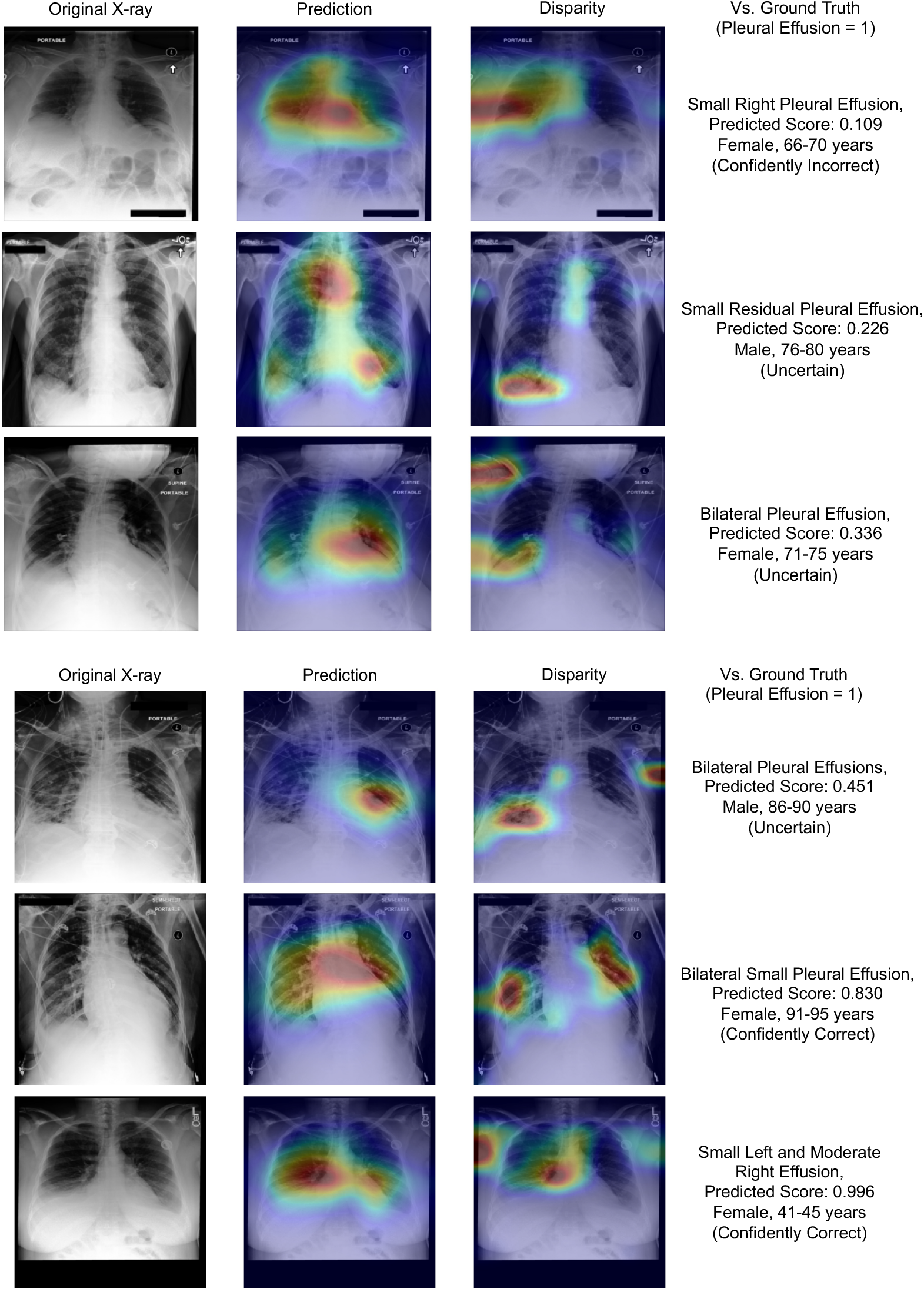
Saliency map examples for the MIMIC-CXR dataset, for the task of detecting pleural effusion. We choose test set examples from the underrepresented subgroup (Black patients) across a range of model predictions. We include the predicted score for effusion, showing the relative model uncertainty. We also include the description of pleural effusion found in the corresponding radiology report. The three columns indicate the original chest X-ray image (Original X-ray), the saliency map for the pleural effusion prediction (Prediction), and the saliency map for the most informative features for error disparity across White and Black patients (Disparity).

### Adapting Models to Underrepresented Subpopulations

Inspired by methods in domain adaptation^34^, we demonstrate how insights from our evaluation procedure can be used to design granular model fine-tuning strategies. For the pleural effusion detection task, we adapt the model to the distribution of Black patients in a given dataset by targeting two distribution shifts that were contributors to the performance disparity: the manifestation shift in image features and the shift in the age distribution. We resample the original training dataset by upweighting the samples corresponding to the image features and age groups that were represented more frequently in the Black population and retrain the final classification layer on the reweighted dataset. We plot the TPR results for the race-adapted model (Fig. 5), showing that our new models achieve greater overall TPR and lower TPR disparity across race with statistical significance (Table S3). We also note that adapting to specific distribution shifts outperforms models that simply employs a race-adaptive threshold for classification. This suggests that our mitigation strategy may be preferable to other approaches as it simply adapts the model at test-time to the distributional composition of patients in a target patient subgroup, instead of potentially compromising overall predictive performance by regularizing or removing information correlated with sensitive attributes in the model prediction.

**Figure 5:**
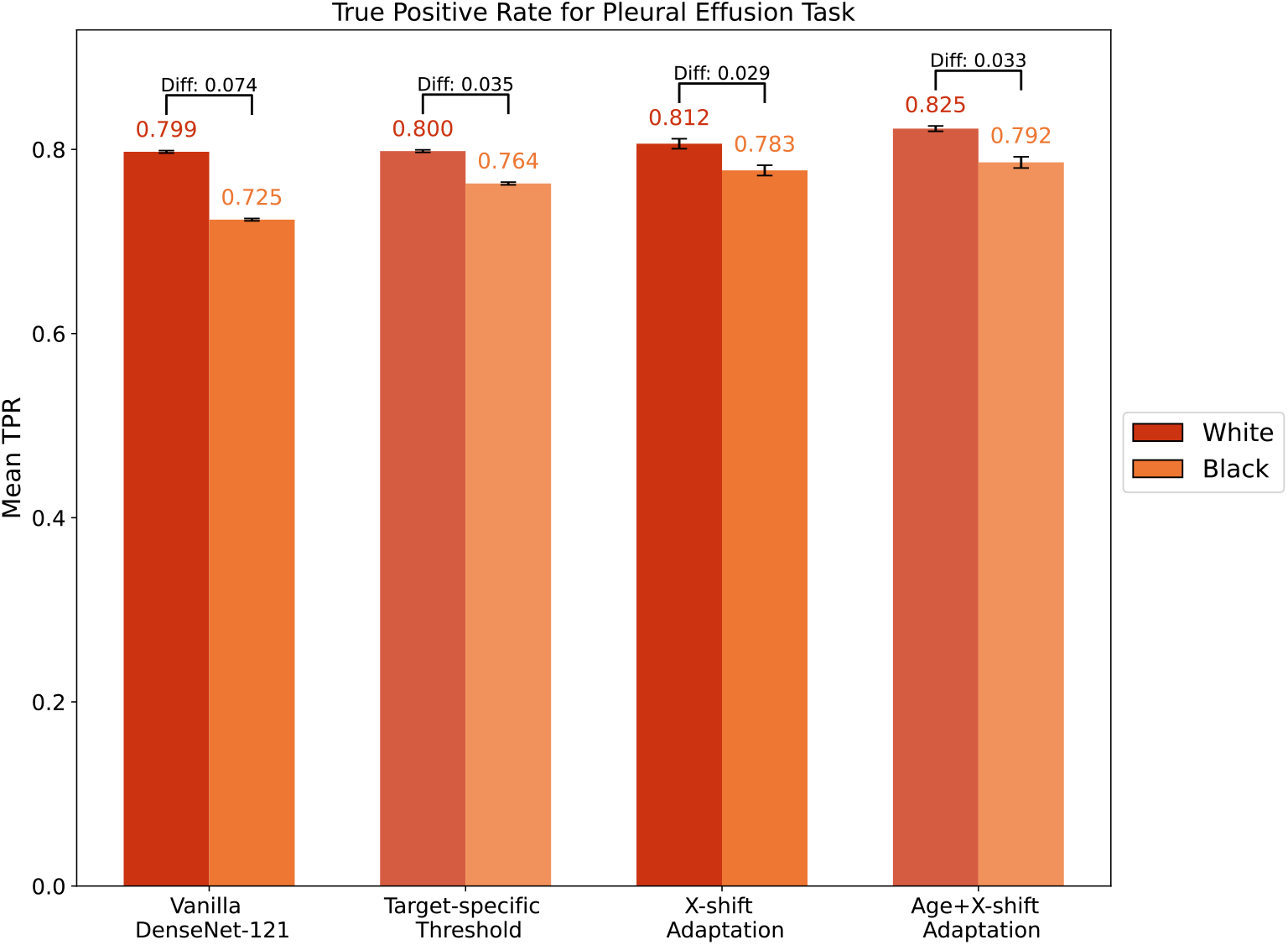
Racial performance disparities for the “Pleural Effusion” task for the DenseNet-121 model in the MIMIC dataset after performing target adaptation, the mean and standard error for TPR is computed for each method. Adapting to shifts in a subset of the image features (X-shift adaptation) increases the TPR for Black patients to 0.783. Additionally adapting to age shifts further increases TPR to 0.792. Interestingly, the detection rates are uniformly higher for the adapted models, showing the benefits of resampling the dataset to improve the representation of specific age groups and image features. We include a baseline (target-specific threshold) which takes the original vanilla DenseNet-121 model and computes an optimal classification threshold for each racial subgroup separately.

## DISCUSSION

In this study, we perform a systematic study to understand why underdiagnosis bias arises between racial subgroups for AI-based chest X-ray disease classification models. We show empirically that the mechanisms causing underdiagnosis bias are surprisingly consistent across datasets, and a nuanced understanding of model and dataset biases are required to interpret subgroup-level diagnostic disparities. Our results for CXR-Foundation indicate that while performance disparities may be partially mitigated with state-of-the-art models, the overall patterns of racial underdiagnosis bias and the mechanisms for causing systematic differences in performances are agnostic to model capacity. Finally, we show that our evaluation method can inform fine-tuning strategies to adapt the model to specific subpopulations.

Our analysis addresses a fundamental limitation of subgroup-level model evaluation, which is increasingly important as large-scale clinical ML models are being evaluated on diverse patient subpopulations. While no prior approach systematically accounts for distributional differences of confounding factors across subpopulations, one commonly employed solution is “slicing” the evaluation dataset to compute model performance on finer subgroups or resampling to balance the representation of confounding variables^4^. However, instead of heuristically choosing which demographic variables to adjust for, we demonstrate that our strategy of jointly modeling ‘error contributions’ can reveal interactions between confounding attributes and provide granular insights through retrospective evaluation of an ML model. We note that our strategy can flexibly incorporate increased granularity of potential confounding attributes and variable groupings depending on the granularity of meta-data recorded in datasets. The significance of collecting meta-data and patient characteristics for image-based models cannot be overstated, as access to granular confounding attributes is necessary to uncover the mechanisms underlying subgroup-level disparities.

The study also offers a cautionary note for practitioners aiming to evaluate or ensure fairness in AI-based medical imaging diagnosis. Well-intentioned strategies for enforcing error rate parity have been proposed to mitigate performance disparities^35^, or to mitigate reliance on spurious attributes or “shortcuts” in the image^36^. However, applying crude adjustment methods without considering implicit distribution shifts have been shown to lead to unintended or harmful consequences. For example, eliminating age-related information in model prediction or model embeddings may be harmful if age is correlated with disease-related characteristics such as the severity, size and location of pleural effusion. Therefore, we stress the importance of disentangling the sources of subpopulation heterogeneity within a target patient population to assess the appropriateness of bias-mitigation strategies. We also propose an alternative mitigation strategy that simply adapts the model to the distributional composition of patients in a target subgroup, instead of removing information correlated with sensitive attributes in the model prediction.

We acknowledge that claiming unfairness in the context of deep learning-based diagnosis of chest X-rays requires further consideration and analysis. We emphasize that our main objective is not to determine whether the deep learning models exhibit unfairness. Various fairness definitions exist in the literature, based on different statistical and causal constraints on model performance metrics^2,37,38^. While our work provides a practical approach for understanding how dataset biases contribute to population-level error rate disparity, understanding fairness violations at increased levels of granularity is generally challenging. For instance, even if the model satisfies equalized error rates across subgroups in a representative and balanced evaluation set, the model may still encode spurious sensitive features in the chest X-ray image to produce individual predictions. However, it is also unclear what constitutes a sensitive feature that should be deemed impermissible and removed, particularly in disease diagnosis. Various works in the fairness literature suggest that the use of characteristics related to subgroup membership may be desired depending on the task and clinical context^39-41^, as disease prevalences in demographic subgroups can act as informative priors.

Our approach is flexible and can incorporate recent advancements made in the pipeline for chest X-ray classification. For example, since our attribution method is agnostic to the base disease detection model, we demonstrate its use with a foundation model that has much larger capacity and is pretrained on completely distinct datasets. In addition to the model itself, other improvements such as leveraging large language models (LLMs) for label and confounder proxy extraction from radiology reports can be incorporated to reduce errors. While label noise in training and evaluating the disease detection model is a limitation in our analysis, our insights regarding dataset biases and our methodology for performing granular subgroup analysis remain applicable as better label curation methods are introduced. Additionally, our method can integrate any improvements in saliency mapping and interpretability methods. Finally, while our analysis focuses on disparities in disease classification performance, our framework can be applied to disparities in alternative task definitions. For example, one may be interested in decomposing the disparities in radiology report generation performance by decomposing an evaluation metric related to the likelihood of the correct diagnosis being generated.

## METHODS

### Attributing Performance Disparity to Observed Distribution Shifts

We use the method from Zhang et.al.^15^ to attribute degradation in model performance to shifts from a set of possible distribution shifts. Multiple distributions can shift simultaneously in practice, and some distribution shifts may have no impact on model performance. To attribute the contribution of a given distribution shift to model performance, the method relies on Shapley values^43^ to attribute changes in model performance to *individual distributions*. The key distinction from existing work in Shapley based feature importance is that the method from Zhang et. al. is directly able to identify the contribution of distributional changes or mechanisms as opposed to the sensitivity to adding/deleting individual features to explain performance changes, in addition to allowing for this attribution in terms of mechanisms or variables not observed by the model, enabling us to attribute performance differences to systemic causes, as opposed to in-vivo model perturbations. A detailed description of the estimation procedure is provided in the Supplement.

### Sensitivity Analysis with Extracted Text-based Proxies

We aim to further decompose the error contribution from the X-ray image features *p*(*X*|*Z*, *D*, *C*) using text-based proxies of unobserved confounders (Fig. 1b). These attributions correspond to the error disparity unexplained by distributional differences in the composition of demographics, pathology, and view-position.

For each chest X-ray with pleural effusion, we perform an automated extraction of pleural effusion size (small, moderate, large) and location (left, right, bilateral) characteristics, as well as radiologist uncertainty from the corresponding radiology report using Radgraph^44^. Each characteristic is represented with a binary variable indicating its presence in the radiology report. For cases where the characteristic is unrecorded, we perform conditional mean imputation by training a DenseNet-121 classifier that predicts the missing characteristic of the effusion. The extracted and predicted values are included as an additional aggregate variable to assess the contribution of pathology manifestation differences to error disparity. We analyze the sensitivity of the remaining attributions when effusion characteristics are included as an upstream variable.

### Interpreting Shifts in Image-level Features using Saliency Maps

Finally, we aim to attribute the error disparity to specific image features that are represented more frequently in Black patients compared to their White patient counterparts. Saliency methods have been validated in chest X-ray disease classification and have been shown to localize regions of interest when the model is able to detect the disease successfully^45^. Using the feature embedding learned by the model, we propose an approach to approximate which regions of the image explain the error disparity resulting from manifestation shift in *p*(*X*|*Z*, *D*, *C*). We define a variable *ϕ*(*X*)_*i*_ for each dimension *i* of the chest X-ray feature embedding, where *ϕ* denotes that the input X-ray has been transformed by the (pretrained) model. We opt for a simple heuristic to detect which variable *ϕ*(*X*)_*i*_ contributed to the error disparity. Given the large dimensionality, we first screen the variables using L1-penalized logistic regression to predict race group from the embedding features. For the selected indices, we introduce the factor *p*(*ϕ*(*X*)_*i*_|*Z*, *D*, *C*) to approximate the manifestation shift in the attribution method and select *i* with the largest error contribution.

To map each of these variables *ϕ*(*𝓍*)_*i*_ to the chest X-ray input image space, we use a Grad-CAM-based saliency method^32,33^. For (x, y)-coordinates in the image space, the saliency map *M*_*0*_(*𝓍*, *y*) is computed for the top-k selected *ϕ*(*𝓍*)_i_ variables, and *∑*_*0*_ *M*_*0*_(*𝓍*, *y*) denotes the sum of the k saliency maps. We select k=3 for our analysis. The saliency method maps the feature embedding to regions in the original image, corresponding to image features that differ across racial groups given the same patient demographics and clinical characteristics.

### Adaptation Strategy for Fine-grained Distribution Shifts

To adapt the model to the target patient distribution (Black patients), we retrain the final linear classification layer of the DenseNet-121 model using a resampled version of the training dataset. We compute the weights for resampling the dataset based on an estimation of the density ratios for two distribution shifts across the entire population and the subgroup of Black patients in the training set. For the X-shift, we use the top-k (k=3) chest X-ray feature embeddings extracted and visualized in the saliency method. We use the same method^2^ to estimate the density ratio 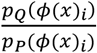, which upweights the samples containing image features that are represented more frequently in the Black population. The weights for the age distribution shift are obtained in the same manner.

### Model Training & Disparity Attribution Experiment Details

The backbone architecture across all experiments for the disease diagnosis model is the 121-layer DenseNet, with the weights initialized using ImageNet pretraining. We applied data augmentation via center crop, random horizontal flip, and 15° random rotation for model training. The optimization is conducted using Adam with default parameters and binary cross-entropy loss. A batch size of 48 is used due to computation limitations. Learning rate is initially set to 0.0005 and drops to half if validation loss does not improve over three epochs. An early stopping condition was implemented if validation loss does not improve over 5 epochs. For binarizing the model prediction, we follow the practice in Seyyed-Kalantari et. al.^5^ and use the single best threshold that maximizes the F1 score in the entire test dataset across all demographic groups for each task.

For the CXR-Foundation^22^ model, we use the 1376-dimensional embeddings which can be obtained by calling the CXR-Foundation API as outlined in https://github.com/Google-Health/imaging-research/tree/master/cxr-foundation. We finetune logistic regression models for the downstream tasks. A batch size of 256 is used. The same optimization parameters are used as the DenseNet-121 model.

Since the dataset contains missing values for the demographic variables of interest, we filter out the subset of the data that has any missing demographic attribute for the analysis of performance disparities. We create an aggregated dataset using the filtered patients that belonged in the original training and validation fold for fitting the domain classifiers for importance weighting. We use logistic regression models throughout for estimating importance weights. The output probabilities from each domain classifier are clipped at 0.99, and each final importance weight is also clipped using a threshold of 99.9 percentile. We perform 100 bootstraps with the aggregated training dataset for the domain classifier training to simulate sampling variability. For each bootstrap, we compute the subgroup-level error metrics based on the fixed prediction probabilities and optimal threshold from the model and compute the attributions using a fixed held-out test set. This ensures that the subgroup-level performance disparity based on the error metric is fixed, but variability is introduced in the attributions through the differences in importance weights computed by the domain classifier at every bootstrap iteration.

## Data Availability

The MIMIC-CXR dataset and Chexpert dataset are publicly available at https://physionet.org/content/mimic-cxr/2.1.0/ and https://stanfordmlgroup.github.io/competitions/chexpert/. Emory-CXR is available upon request to the authors due to privacy protection.

---

**Extended Data Fig. 1.**
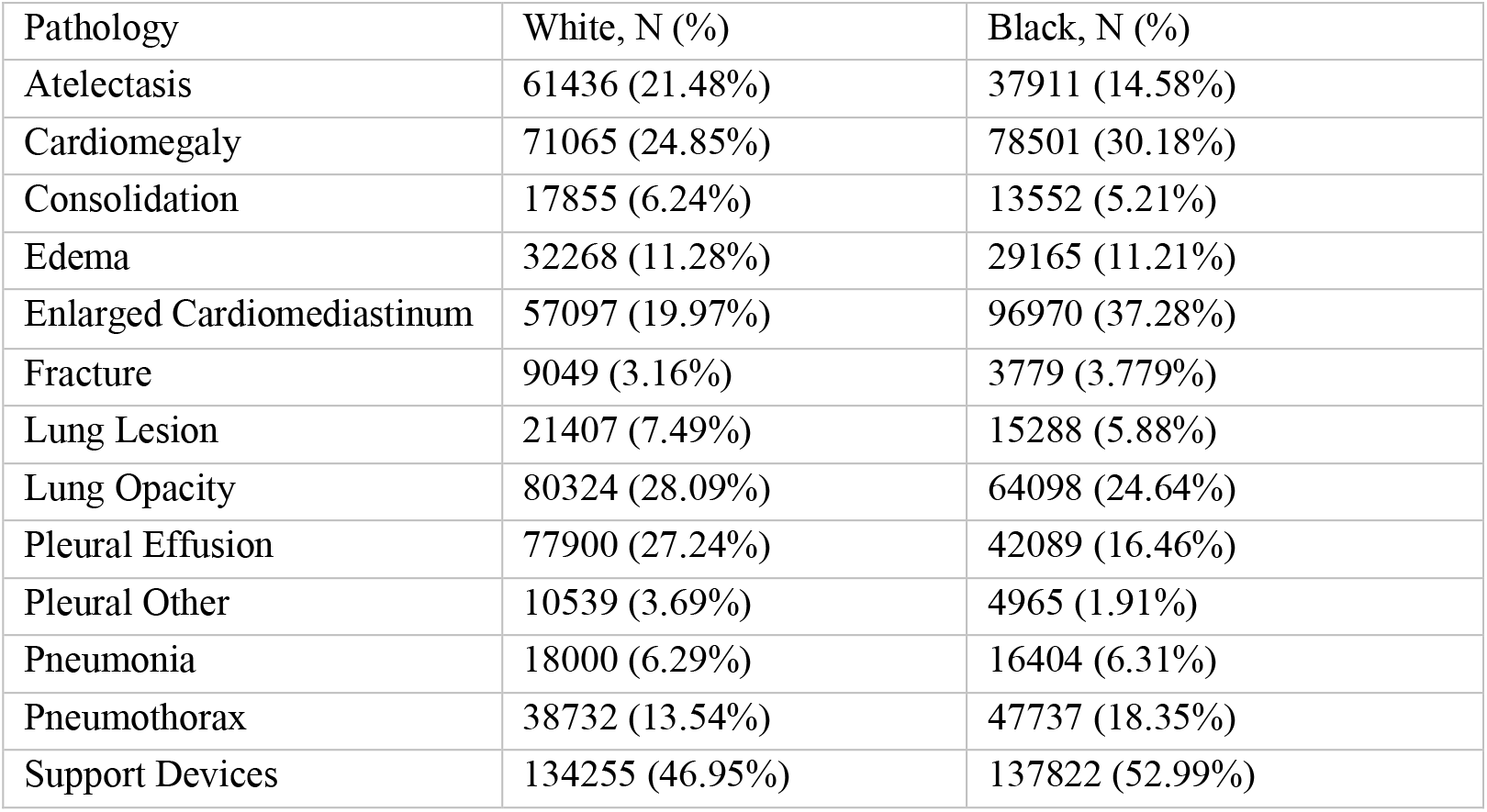
Prevalence of pathologies across racial groups (Emory)

**Extended Data Fig. 2.**
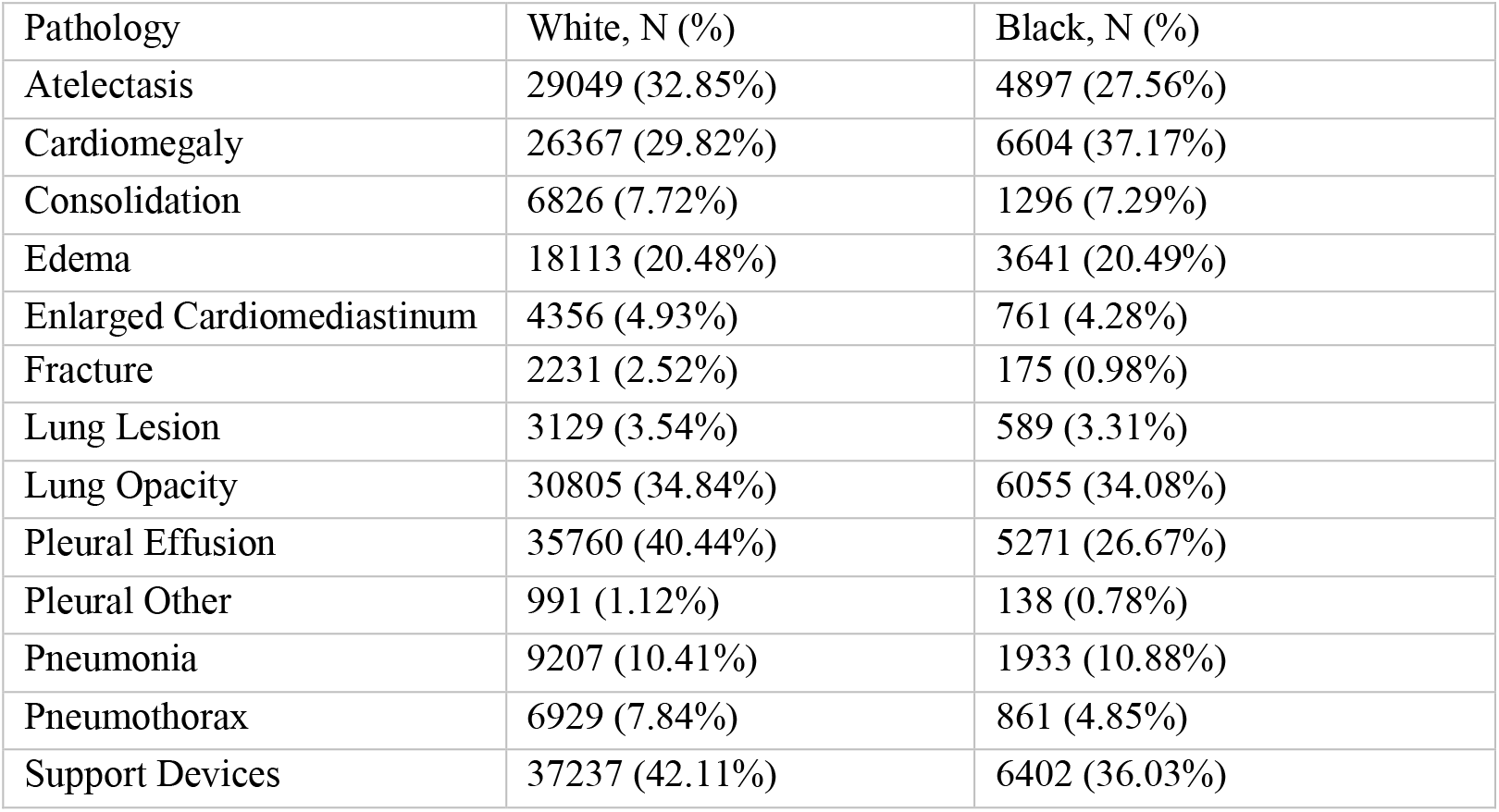
Prevalence of pathologies across racial groups (MIMIC)

**Extended Data Fig. 3.**
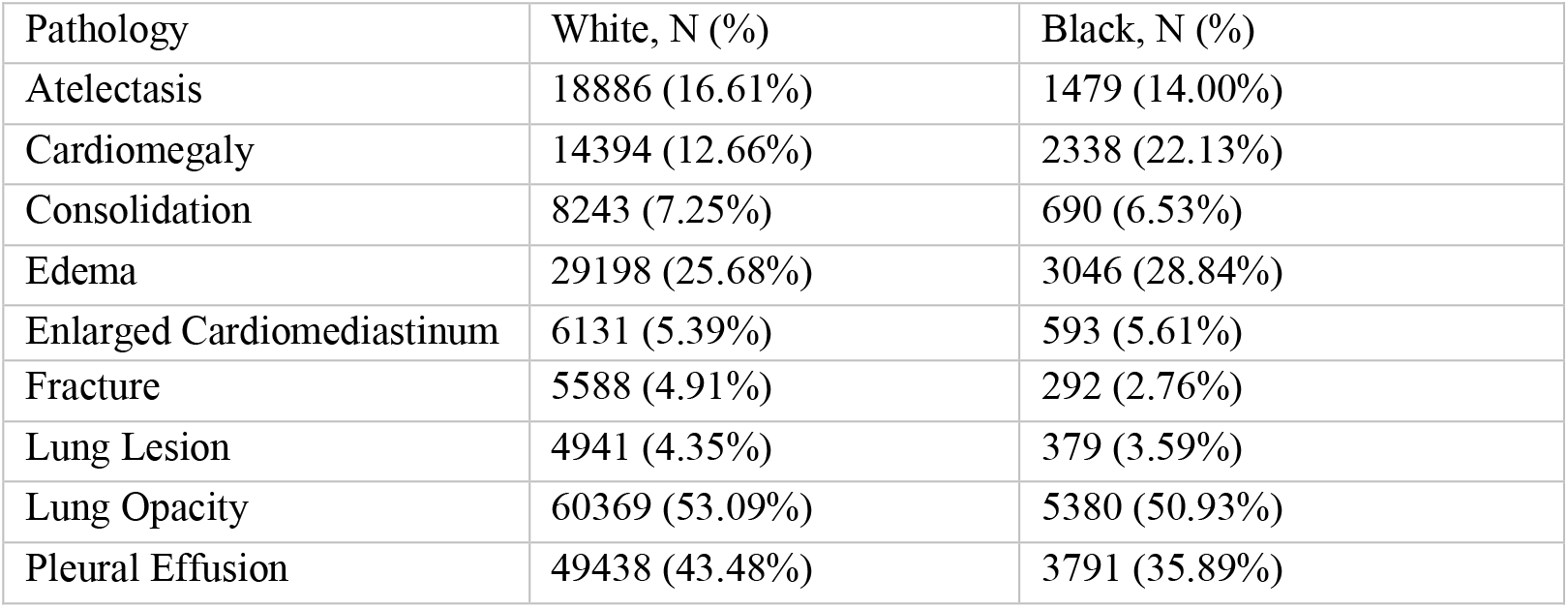
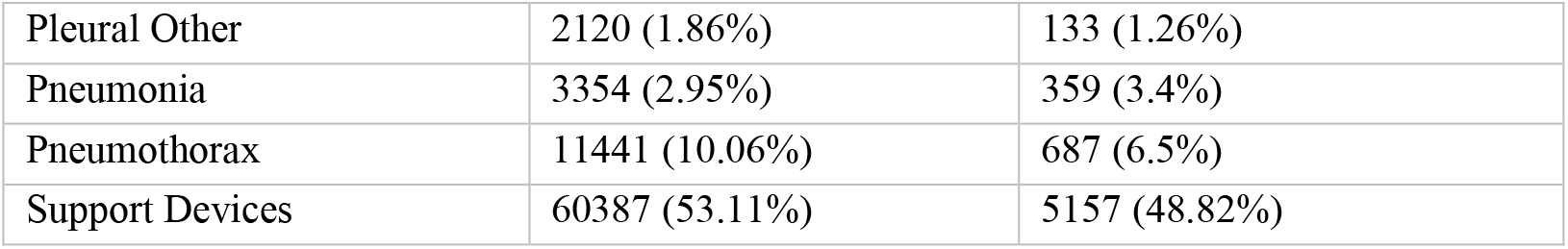
Prevalence of pathologies across racial groups (CheXpert)

**Extended Data Fig. 4.**
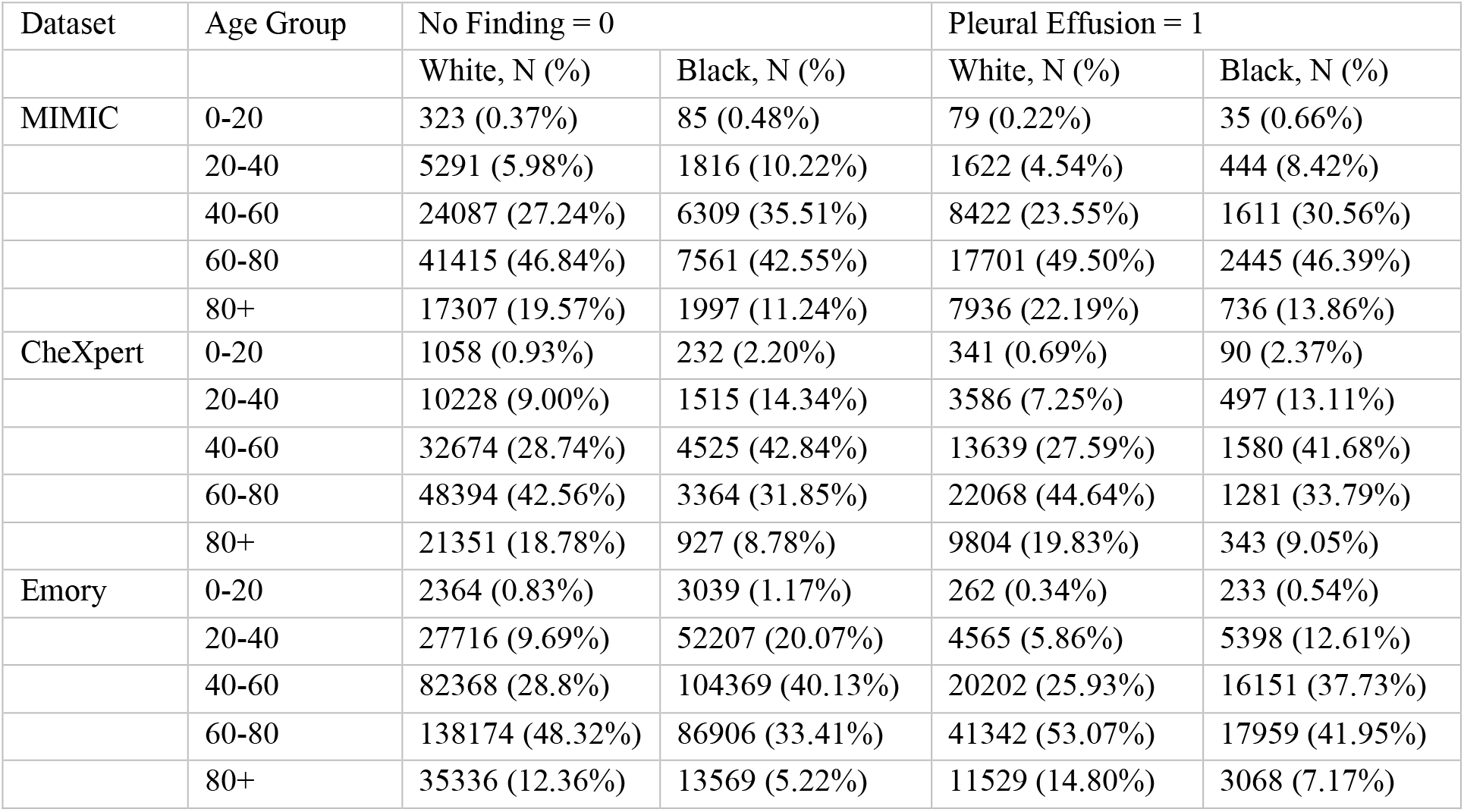
Distributional differences of age across racial groups, for patients with No Finding = 0 and for patients with Pleural Effusion = 1.

