## Supplementary Materials for "Underdiagnosis Bias of Chest Radiograph Diagnostic AI can be Decomposed and Mitigated via Dataset Bias Attributions"

**Table S1.** Model Performances Comparison Across Models, No Finding task. For the race comparison, we compare the distribution of FPRs obtained from the DenseNet-121 model in each race group to the baseline (White patients) model performance within the same dataset. For model comparison, we compare the distribution of model performances across models for each race group. P-values are obtained using the Wilcoxon rank-sum test. FPR distributions are obtained using 20 bootstrap iterations.

| Dataset | Race Group | DenseNet-121 | CXR-Foundation | Race Comparison | Model Comparison |
|---|---|---|---|---|---|
| | | Mean FPR (SD) | Mean FPR (SD) | p-value | p-value |
| MIMIC | White | 0.131 (0.006) | 0.118 (0.006) | N/A | <0.001 |
| | Black | 0.193 (0.009) | 0.173 (0.009) | <0.001 | <0.001 |
| | Hispanic | 0.219 (0.006) | 0.202 (0.008) | <0.001 | <0.001 |
| | Asian | 0.141 (0.005) | 0.142 (0.008) | <0.001 | 0.725 |
| CheXpert | White | 0.087 (0.009) | 0.078 (0.006) | N/A | 0.003 |
| | Black | 0.112 (0.011) | 0.105 (0.008) | <0.001 | 0.022 |
| | Hispanic | 0.148 (0.012) | 0.133 (0.01) | <0.001 | 0.001 |
| | Asian | 0.100 (0.009) | 0.093 (0.008) | <0.001 | 0.02 |
| Emory | White | 0.269 (0.009) | N/A | <0.001 | N/A |
| | Black | 0.250 (0.008) | N/A | <0.001 | N/A |
| | Hispanic | 0.348 (0.013) | N/A | <0.001 | N/A |
| | Asian | 0.293 (0.01) | N/A | <0.001 | N/A |

**Table S2.** Model Performances Comparison Across Models, Pleural Effusion task. For the race comparison, we compare the distribution of TPRs obtained from the DenseNet-121 model in each race group to the baseline (White patients) model performance within the same dataset. For model comparison, we compare the distribution of model performances across models for each race group. P-values are obtained using the Wilcoxon rank-sum test. TPR distributions are obtained using 20 bootstrap iterations.

| Dataset | Race Group | DenseNet-121 | CXR-Foundation | Race Comparison | Model Comparison |
|---|---|---|---|---|---|
| | | Mean TPR (SD) | Mean TPR (SD) | p-value | p-value |
| MIMIC | White | 0.797 (0.006) | 0.799 (0.012) | N/A | 0.957 |
| | Black | 0.724 (0.006) | 0.712 (0.016) | <0.001 | 0.006 |
| | Hispanic | 0.811 (0.009) | 0.804 (0.012) | <0.001 | 0.047 |
| | Asian | 0.784 (0.006) | 0.796 (0.013) | <0.001 | <0.001 |
| CheXpert | White | 0.843 (0.006) | 0.821 (0.015) | N/A | <0.001 |
| | Black | 0.782 (0.009) | 0.756 (0.019) | <0.001 | <0.001 |
| | Hispanic | 0.819 (0.006) | 0.793 (0.016) | <0.001 | <0.001 |
| | Asian | 0.868 (0.007) | 0.841 (.016) | <0.001 | <0.001 |
| Emory | White | 0.747 (0.014) | N/A | <0.001 | N/A |
| | Black | 0.686 (0.017) | N/A | <0.001 | N/A |
| | Hispanic | 0.771 (0.024) | N/A | 0.003 | N/A |
| | Asian | 0.763 (0.016) | N/A | 0.005 | N/A |

**Table S3.** Model Performance Comparison Across Adapted Models, Pleural Effusion task. For the model comparisons, we compare the distribution of TPR obtained for the evaluation cohort of each model to (1.) the baseline DenseNet-121 model and (2.) the race-specific threshold model. P-values are obtained using the Wilcoxon rank-sum test. TPR distributions are obtained using 20 bootstrap iterations. We also show the task AUROC for Black patients after adaptation, which shows a slight deterioration for the adapted models.

| Dataset | Model | Mean target AUROC (SD) | Race Group | Mean TPR (SE) | Comparison 1 (P-value) | Comparison 2 (P-value) |
|---|---|---|---|---|---|---|
| MIMIC | DenseNet-121 | 0.8718 (0.00028) | White | 0.7974 (0.0014) | N/A | N/A |
| | | | Black | 0.7237 (0.0013) | N/A | N/A |
| | Race Threshold | 0.8718 (0.00028) | White | 0.7990 (0.0018) | 0.579 | N/A |
| | | | Black | 0.7629 (0.0015) | <0.001 | N/A |
| | Adaptation (X) | 0.8682 (0.00058) | White | 0.8284 (0.0041) | <0.001 | <0.001 |
| | | | Black | 0.7800 (0.0045) | <0.001 | 0.003 |
| | Adaptation (Age&X) | 0.8679 (0.00053) | White | 0.8256 (0.0031) | <0.001 | <0.001 |
| | | | Black | 0.7980 (0.0054) | <0.001 | <0.001 |

**Figure S1.** Sensitivity analysis results using text-based proxies obtained using Radgraph, for pleural effusion task across MIMIC and CheXpert dataset. We use pleural effusion size (small, moderate, large) and location (left, right, bilateral) characteristics, as well as radiologist uncertainty from the corresponding radiology report extracted using Radgraph. The reduced age attribution indicate that the effusion manifestation and age are correlated and jointly explains the performance disparity.  This indicates that the model may be influenced by a combination of purely spurious age-related features, as well as "mixed" features that mix the label signal (pleural effusion) and age, such as effusion size and severity.

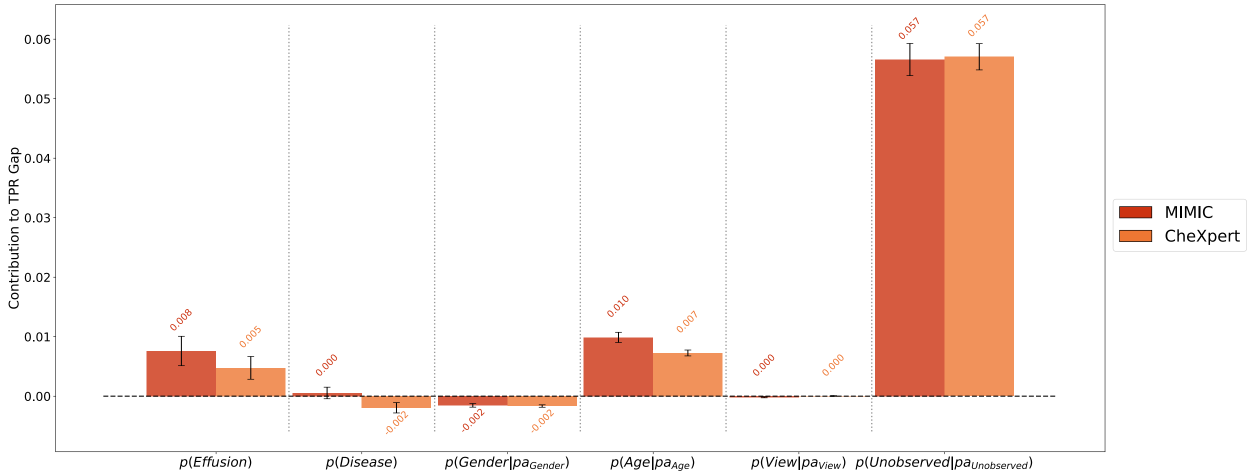

**Figure S2.** Distribution of predicted probabilities for the DenseNet-121 model, for cases where the label indicates that there is no pathology in the X-ray (No Finding = 0). Density is interpolated using kernel density estimation (KDE). Higher prediction probabilities (>0.8) may indicate label error, given that the model is confident that there is a pathology in the X-ray. This suggests that MIMIC-CXR and Emory contain more false positive samples that are likely to be due to labeler error, which may be a source of (false) underdiagnosis bias in the "No Finding" not explained by observed attributes (pathologies, demographics, view position).

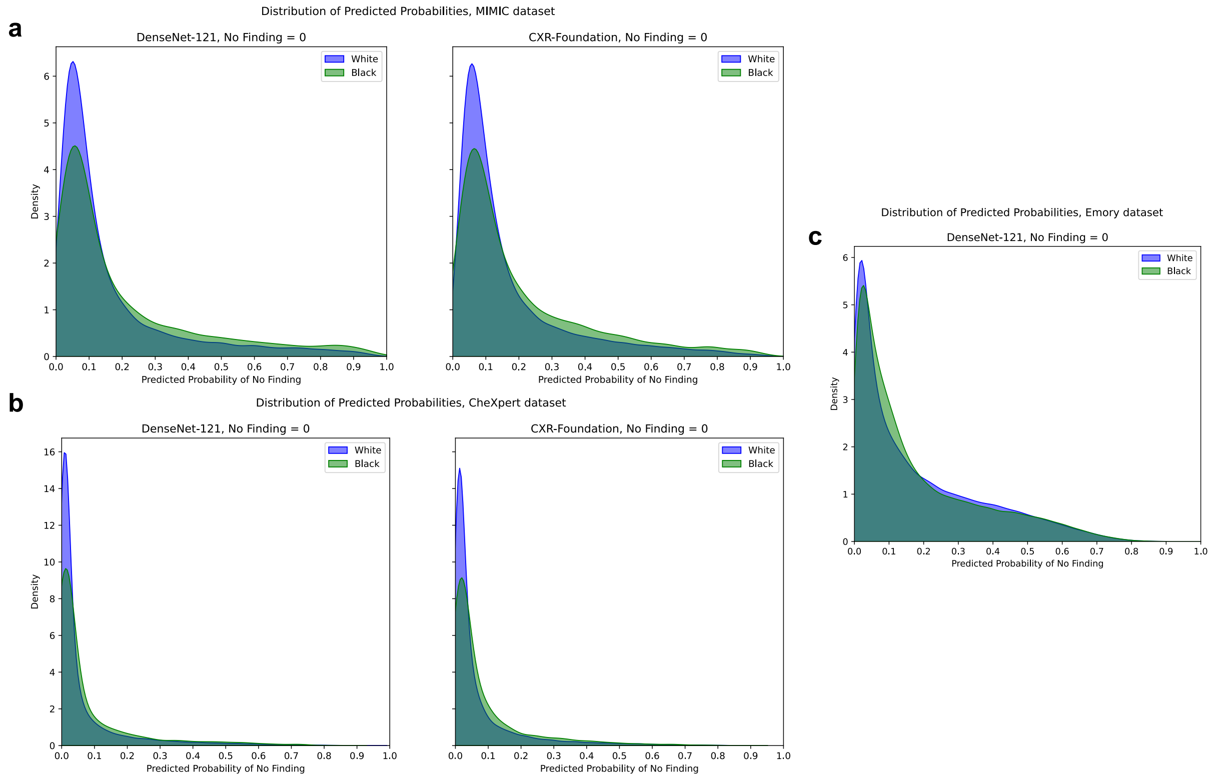

**Figure S3.** Distribution of predicted probabilities for both models, for cases where the label indicates that there is pleural effusion in the X-ray (Pleural Effusion = 1). Density is interpolated using kernel density estimation (KDE). Model is underestimating pleural effusion probability for younger patients.

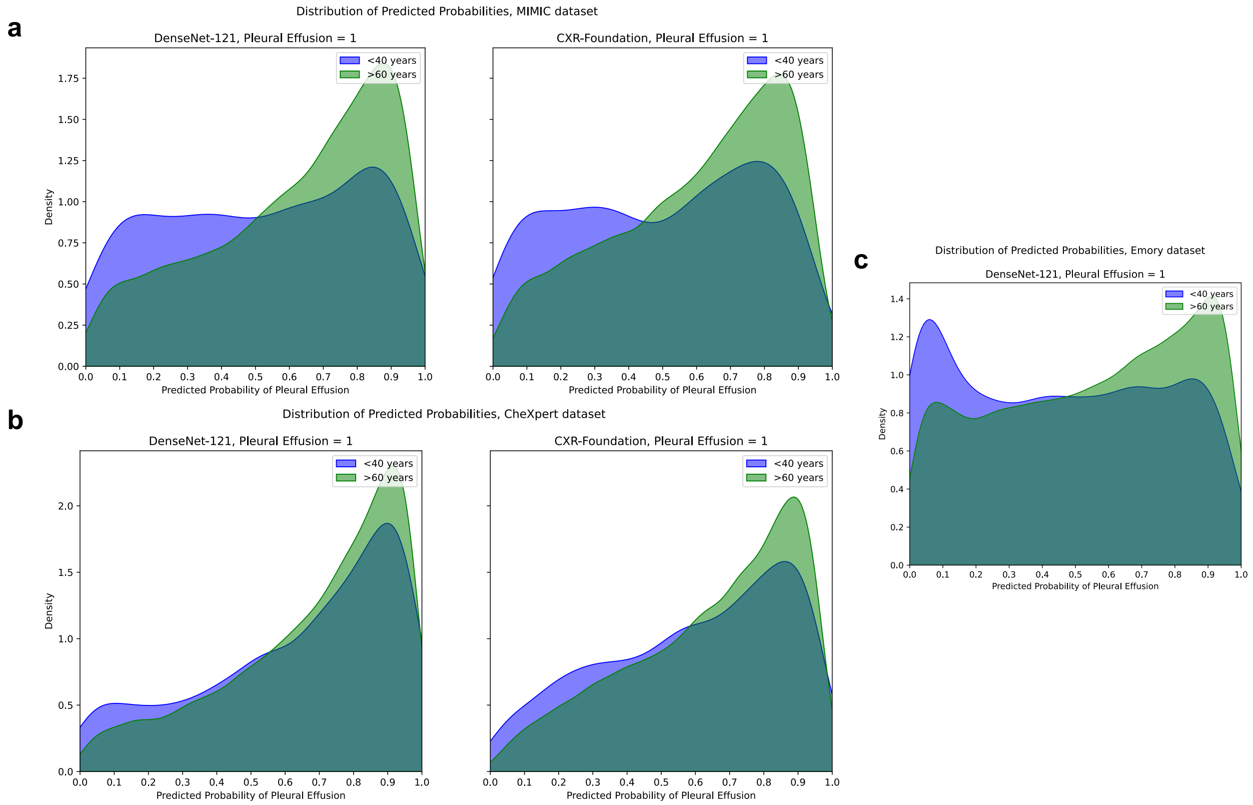

**Causal Models & Diagrams**

A causal model is a set of mechanistic assumptions describing the underlying data generating process for a set of variables under investigation and enables reasoning about the effects of variables on an outcome of interest. In particular, the value of each measured/observed variable is assumed to be determined by an independent mechanism given their direct causes and exogenous noise accounting for factors outside of the model. A causal model induces different types of probability distributions over the set of variables under investigation. One important type of distribution is the observational distribution, which represents the joint data distribution of retrospective or observational data. A causal model embodies an important assumption, which states that every variable is conditionally independent of its non-effects given its direct causes. Therefore, the joint data distributions over the set of observed variables $V$ can be factorized as a product of independent conditional mechanisms, where $pa(V_i)$ represents the causal parents of $V_i \in V$:

$$P(V) = \prod_{i=1}^{N} P(V_i | pa(V_i))$$

Causal diagrams provide a way to visualize the assumptions from a causal model, where each node represents an observed variable and directed edges between variables indicate postulated direct causal influence. A bidirected edge between two variables indicates that there is some shared unobserved information affecting how both variables obtain their values, thus inducing correlation. Note that we do not use the causal model to model interventional distributions, but only to characterize the changes in the joint distribution of variables with respect to a sensitive attribute.

We consider the following causal diagram representing the data-generating process of chest X-rays, where we have binary pathology labels $Z$, demographics $D$, the view position of the x-ray $C$, and the model loss ($L \in \{0, 1\}$ indicating whether the model classified the sample correctly or incorrectly). For the analysis of performance disparities, we treat the X-ray as an unobserved variable (denoted by the unfilled node). While we have access to the representation of the X-ray image via the learned feature embedding, modeling the entire joint distribution is still intractable.

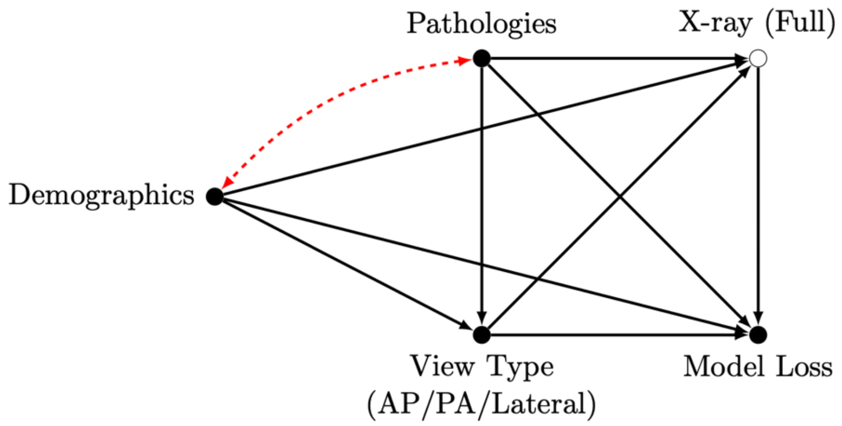

**Method for Computing Attributions Based on Causal Diagram** For a more comprehensive description of the method, we refer readers to Zhang et.al.[1]

We consider a source and target distribution, denoted by $P$ and $Q$ respectively. For example, $P$ and $Q$ can be defined as two racial subgroups. Note that the expectation of the model loss $L$ for each subgroup can be defined over the joint distribution of the observed variables as follows.

$$\mathbb{E}_P[L] = \sum_{z,d,c} \mathbb{E}_P[l \mid z, d, c]\, p_P(z, d, c)$$

$$\mathbb{E}_Q[L] = \sum_{z,d,c} \mathbb{E}_Q[l \mid z, d, c]\, p_Q(z, d, c)$$

For example, using $\mathbb{I}[\cdot]$ as the indicator function, the FPR can be written as

$$FPR = \mathbb{E}_P\big[\mathbb{I}[\hat{Y} = 1] \mid Y = 0\big] = \sum_{z,d,c} \mathbb{E}_P\big[\mathbb{I}[\hat{Y} = 1] \mid z, d, c\big]\, p_P(z, d, c \mid Y = 0)$$

The gap in model loss across subgroups can be written as follows

$$\mathbb{E}_Q[L] - \mathbb{E}_P[L] = \mathbb{E}_Q[L] - \mathbb{E}_S[L] + \mathbb{E}_S[L] - \mathbb{E}_P[L] = \psi_2 + \psi_1$$

$$\mathbb{E}_S[L] = \sum_{z,d,c} \mathbb{E}_P[l \mid z, d, c]\, p_Q(z, d, c)$$

Note that $\mathbb{E}_S[L]$ is a fixed value based on the subgroup definitions. We begin by focusing on the difference $\psi_1 = \mathbb{E}_S[L] - \mathbb{E}_P[L]$, defined as follows.

$$\psi_1 = \mathbb{E}_S[L] - \mathbb{E}_P[L] = \sum_{z,d,c} \mathbb{E}_P[l \mid z, d, c]\, p_Q(z, d, c) - \sum_{z,d,c} \mathbb{E}_P[l \mid z, d, c]\, p_P(z, d, c)$$

To further attribute $\psi_1$ to individual distribution shifts, we simulate expectations of our metric of interest under a hypothetical distribution $\tilde{P}(Z, D, C)$, which is a product of mixed factors from both $P$ and $Q$. Recall the canonical causal diagram encodes a factorization of the joint probability distribution $p(Z, D, C)$, as shown below.

$$p_P(Z, D, C) = p_P(Z) p_P(D|Z) p_P(C|Z, D)$$

$$p_Q(Z, D, C) = p_Q(Z) p_Q(D|Z) p_Q(C|Z, D)$$

Each factor represents a probabilistic mechanism or dependence structure between variables that can shift across racial subgroups. In other words, $\tilde{P}$ corresponds to a joint distribution where only a subset of the conditional mechanisms in the causal factorization have shifted. For example, suppose we would like to quantify the contribution of the shift in disease prevalence $p(z)$, so $\tilde{P} = p_Q(z)p_P(d|z)p_P(c|d,z)$:

$$\mathbb{E}_{\tilde{P}}[L] - \mathbb{E}_P[L] = \sum_{z,d,c} \mathbb{E}_P[l \mid z,d,c]\, p_Q(z)p_P(d|z)p_P(c|d,z) - \sum_{z,d,c} \mathbb{E}_P[l \mid z,d,c]\, p_P(z)p_P(d|z)p_P(c|d,z)$$

We require the following positivity assumption for estimating the quantity $\mathbb{E}_{\tilde{P}}[L]$.

(*Positivity*) For all $z, d, c$ such that $p_Q(z,d,c) > 0$, it must be that $p_P(z,d,c) > 0$, and vice versa.

Let $C$ be the index set of all possible distributions that may shift and suppose $i$ is the index for the distribution in the factorization. So $p^i$ indicates the factors that are shifted to form the coalition $\tilde{C} \subseteq C$. The simulated performance disparity $Val(\tilde{C})$ is given as follows.

$$\begin{aligned} Val(\tilde{C}) &= E_{\tilde{P}}\,[l] \; - \; E_P\,[l] \\ &= E_P\,[w_{\tilde{C}} \cdot \; l] \; - E_P\,[l] \end{aligned}$$

Note the second term can be computed directly from the observed data from $P$. For the first term, the importance sampling weights $w_{\tilde{C}}$ for a given sample $(z,d,c)$ is computed as follows,

$$w_{\tilde{C}}\,(z,d,c) = \frac{p_{\tilde{P}}(z,d,c)}{p_P(z,d,c)} = \prod_{i \in \tilde{C}} \frac{p_Q^i}{p_P^i}$$

We estimate these individual weights by training domain classifiers[2]. Finally, we use the obtained estimations of $Val(\tilde{C})$ to compute the Shapley value attribution for each conditional mechanism $p_i$

$$Attr(p_i) = \frac{1}{|C|} \sum_{\tilde{C} \subseteq C \setminus \{p_i\}} \binom{|C|-1}{|C|}^{-1} \left(Val(\tilde{C} \cup \{p_i\}) - Val(\tilde{C})\right)$$

**Connection to Shortcut Learning & Mitigation.** A shortcut variable is a variable spuriously (as opposed to meaningfully) correlated with the target variable, and the association may differ by subgroups. A ML model that learns to predict using this association in one subgroup may not generalize to subgroups where the association differs. Various works have formalized the concept of shortcut learning though a causal framework[4,5], and some works have applied these concepts to explain performance disparities in chest X-ray disease classification[6,7]. Note that input features can be composed of features that are (noisy) functions of only the shortcut variable (completely spurious)[4], or features that mix the target label and spurious feature[5].

We ground our explanation using a concrete example, with pleural effusion as the target label and age as the shortcut variable. Our results indicate that for age, the X-ray includes features that are completely spurious features (ex. bone density), as well as features that mix age and pleural effusion (ex. size and severity of effusion may be influenced by age). Therefore, crude approaches of removing age-based shortcuts may have unintended consequences for model performance and generalization.

Let $y$ be the target label (pleural effusion), $x^*$ be the disease-related features of the x-ray (encoding disease severity), $x$ be the full x-ray, and $z$ be the shortcut variable (age). Given a model $f = \hat{p}(y|x)$, the expected loss can be computed as follows

$$\mathbb{E}_f[l] = \sum_{y,x^*,x,z} \mathbb{E}_f[l \,|y, x^*, x, z]\, p(y, x^*, x, z)$$

$$\mathbb{E}_f[l] = \sum_{y,x^*,x,z} \mathbb{E}_f[l \,|y, x^*, x, z]\, p(y) p_D(z|y) p(\, x^*, x|y, z)$$

Note that $p_D(z|y)$ represents the correlative relationship between the shortcut variable and the target label. While the strength of this correlation can be varied, a common choice of $p_D(z|y)$ used to detect shortcut learning in previous works is $p(z)$. This equates to removing the bidirected edge between these nodes in the causal graph, for example between age and pleural effusion.

Suppose we have the scenario where the disease-related features $x^*$ is generated using functions that mix the target label and shortcut variable. We represent this scenario using the following SCM with a parameter $a$:

$$z \leftarrow \mathcal{N}(ay + 0.5, 1)$$
$$y \leftarrow Bern(0.5)$$
$$x^* \leftarrow \mathcal{N}((0.3 + 0.2z)y - 0.5, 1)$$

In this setting, the correlation strength between $x^*$ and $y$ depends on $z$, and the spurious correlation strength between $y$ and $z$ depends on the parameter $a$. For example, different racial groups may have different values of $a$. Suppose for White patients we have $a = 0$. Under this SCM, there is no correlation between age and effusion, and the relationship between $x^*$ and $y$ is given by

$$x^* \leftarrow \mathcal{N}(0.4y - 0.5, 1)$$

However, suppose for Black patients we have $a = 1$, now the relationship between $x^*$ and $y$ is given by

$$x^* \leftarrow \mathcal{N}(0.5y - 0.5, 1)$$

Intuitively, a model will learn a fixed relationship between pleural effusion severity in the X-ray image and the label for pleural effusion under a given age distribution. By changing the spurious relationship between age and the target label in the training data distribution, the model may encode a different relationship between effusion severity and the effusion label.

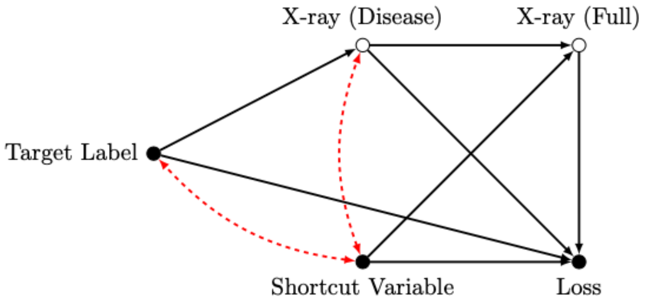


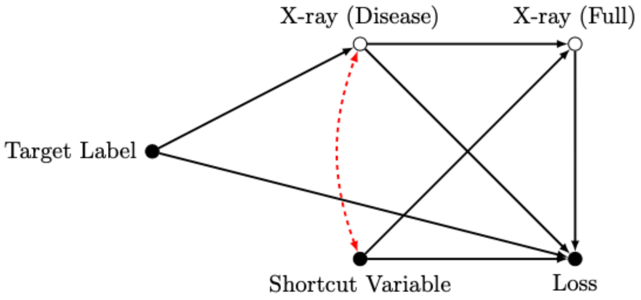


Causal graph before and after crude approach of decorrelating the shortcut variable and the target label (setting $a = 0$ in the SCM). A dependence still exists between the shortcut variable and target through the disease-related X-ray features via mixing.
